# ViralFlow: an automated workflow for SARS-CoV-2 genome assembly, lineage assignment, mutations and intrahost variants detection

**DOI:** 10.1101/2021.10.01.21264424

**Authors:** Filipe Zimmer Dezordi, Túlio de Lima Campos, Pedro Miguel Carneiro Jeronimo, Cleber Furtado Aksenen, Suzana Porto Almeida, Gabriel Luz Wallau, Fiocruz COVID-19 Genomic Surveillance Network

**Affiliations:** Instituto Aggeu Magalhães, Departamento de Entomologia e Núcleo de Bioinformática, Fiocruz, Campus da UFPE - Av. Prof. Moraes Rego s/n, 50670-420, Recife, Brazil; Instituto Aggeu Magalhães, Núcleo de Bioinformática, Fiocruz, Campus da UFPE - Av. Prof. Moraes Rego s/n, 50670-420, Recife, Brazil; Oswaldo Cruz Foundation (Fiocruz), Branch Ceará, Eusebio 61760-000, Fortaleza, Ceará, Brazil

## Abstract

The COVID-19 pandemic, a disease caused by the Severe Acute Respiratory Syndrome coronavirus 2 (SARS-CoV-2), emerged in 2019 and quickly spread worldwide. Genomic surveillance has become the gold standard methodology to monitor and study this emerging virus. The current deluge of SARS-CoV-2 genomic data being generated worldwide has put additional pressure on the urgent need for streamlined bioinformatics workflows for data analysis. Here, we describe a workflow developed by our group to process and analyze large-scale SARS-CoV-2 Illumina amplicon sequencing data. This workflow automates all the steps involved in SARS-CoV-2 genomic analysis: data processing, genome assembly, PANGO lineage assignment, mutation analysis and the screening of intrahost variants. The workflow presented here (https://github.com/dezordi/ViralFlow) is available through Docker or Singularity images, allowing implementation in laptops for small scale analyses or in high processing capacity servers or clusters. Moreover, the low requirements for memory and CPU cores makes it a versatile tool for SARS-CoV-2 genomic analysis.

## INTRODUCTION

The emergence (Wu et al., 2020) and rapid spread of the Severe acute respiratory syndrome coronavirus 2 (SARS-CoV-2), the virus which causes the Coronavirus Disease 2019 (COVID-19), and the subsequent establishment of the COVID-19 pandemic (WHO, 2020), triggered a global effort to sequence and identify the circulating SARS-CoV-2 lineages. This effort resulted in the rapid availability of more than 2 million genomes into the EpiCoV™ database hosted on GISAID in July 2021 (Shu & McCauley, 2017), representing more than 1,700 lineages described on PANGO lineages (O’Toole et al., 2021).

A range of molecular biology methods have been developed to diagnose SARS-CoV-2 infections, such as RT-qPCRs, RT-LAMP, immunoassays or Sanger sequencing (Bezerra et al., 2021; da Silva et al., 2020; Nörz et al., 2021). However, only whole genome sequencing can provide enough genetic information for reliable lineage discrimination and characterization of genome wide mutation patterns that are necessary for the characterization of variants of concern (VOCs) (Lauring & Hodcroft, 2021). Amplicon-based Next generation sequencing (NGS) has become the gold standard methodology for SARS-CoV-2 genome sequencing (Charre et al., 2020), but such abundance of sequencing data from hundreds/thousands of samples also brings new challenges at bioinformatic analysis. At the moment, the Centers for Disease Control and Prevention (CDC) official git repository contains eighteen sources of bioinformatics tools to deal with different SARS-CoV-2 sequencing data (SARS-CoV-2 Sequencing Resources, 2020). However, a single validated workflow that is able to perform several key genomic analyses such as: data quality check, genome assembly, virus lineage assignment, mutation description and intrahost variants variability analysis by sample is still lacking.

In this work, we describe a workflow currently in use by the Fiocruz COVID-19 Genomic Surveillance Network (Naveca et al., 2021; Paiva et al., 2020; Resende et al., 2021). It was developed to work with paired-end Illumina amplicon sequencing reads, and is focused on both pre- and post-genomic analysis. It was designed to attend research groups with diverse computational structures such as: personal computers and multi-user servers through the containerization of the workflow with Docker (https://www.docker.com/) and Singularity (https://sylabs.io/singularity/).

## MATERIAL AND METHODS

### Workflow structure

The workflow was developed within an Ubuntu 20.04.2 LTS Docker environment (https://hub.docker.com/_/ubuntu) and is composed of five major steps to analyse SARS-CoV-2 Illumina paired-end amplicon sequencing data (**Figure 1a**): quality control; consensus generation; intrahost variant analysis; virus lineage assignment and mutation analysis; and Assembly metrics analytics, and can be used in different computational environments (**Figure 1b**).

**Figure 1.**
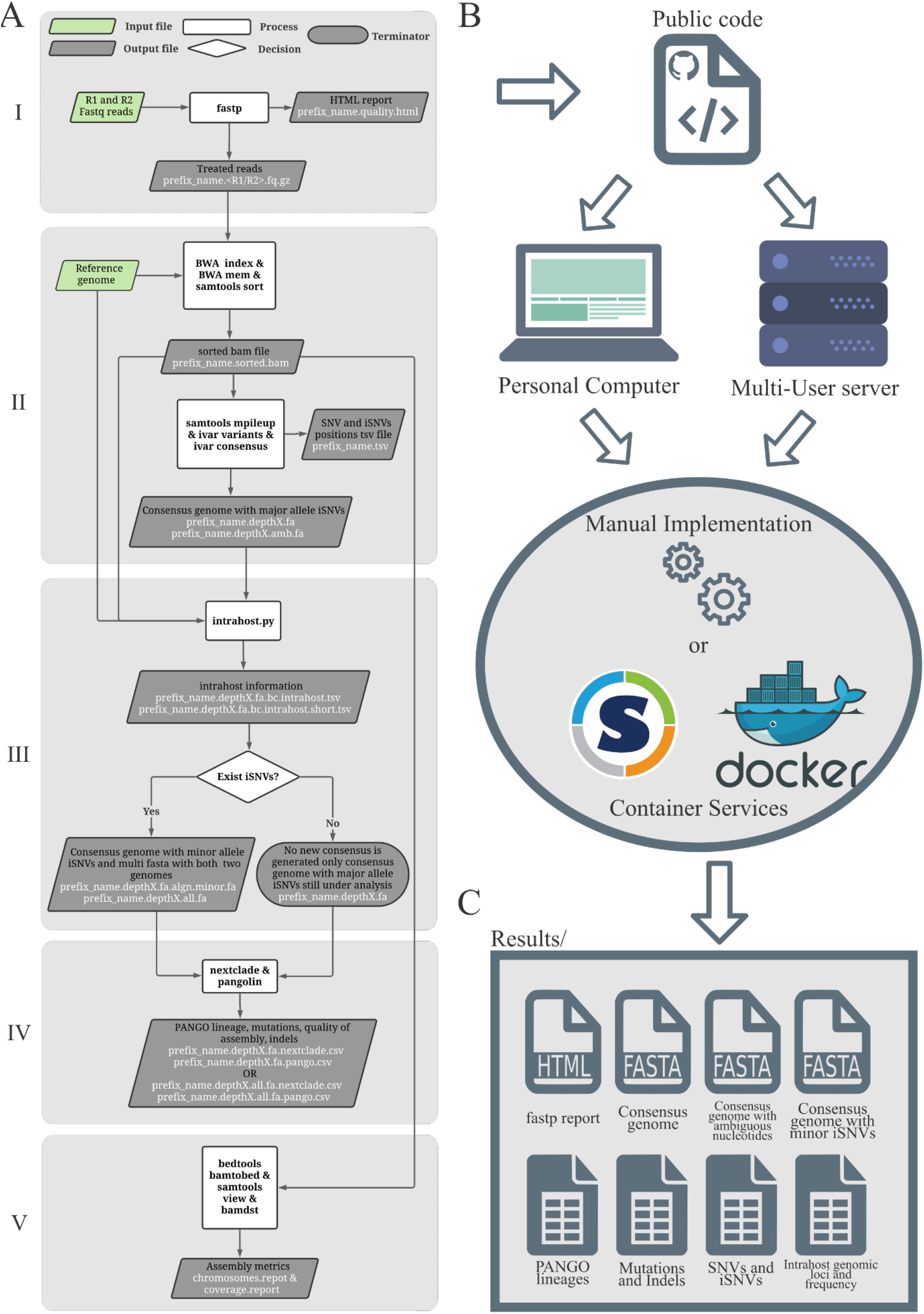
Workflow scheme. **a**. The workflow structure with the five steps. **b**. The workflow can be configured to work on diverse computational environments. **c**. Several outputs generated by the workflow per sample.

The quality control step is performed with fastp v.0.20.1 tool (Chen et al., 2018), where the paired-end reads are trimmed using a minimum read quality threshold of Phred score 20. The adapters or the PCR primers and the minimum length threshold for trimmed reads should be parsed by the user. In addition to the paired-end treated data, this step provides an html with information of pre and post-treatment steps (**Figure 1c**).

The generation of the consensus genome is performed using the reference-guided assembly strategy. In this step a mapping of paired-end libraries against a reference genome is performed with BWA v.0.7.17 (Li & Durbin, 2009), we recommend the SARS-CoV-2 reference genome Wuhan-Hu-1, NCBI refseq NC_045512.2 code. The Samtools v.1.9 (Li et al., 2009) and iVar v.1.3.1 (Grubaugh et al., 2019) tools are used to correct recovery the SNPs and indels were two consensus are generated: one with the majority allele present into every nucleotide position along the genome (ivar consensus -t 0) and another version with ambiguous nucleotide characters, in cases where majority intrahost Single Nucleotide Variants (iSNVs) encompass up to 60% of allele frequencies (ivar consensus -t 0.6). Only mapped bases with quality equal to or greater than 30 (-q 30) were used into iVar counts. Minimum depth threshold to consider a position with supported intrahost variants should be parsed by the user.

Considering that iVar did not provide an option to generate consensus harboring all iSNVs with two or more alleles found in low frequency - and the consensus genome with minor iSNVs is essential to understand the effect of intrahost variants - we developed an *in house* python script (intrahost.py) which uses the allele frequencies per position output of bamreadcount v.0.8.0 (Khanna et al., 2021) to recognize only the positions with 2 or more alleles and generate a consensus harboring all minor supported alleles. To avoid the recognition of sequencing artifacts such as intrahost variants signals, genomic positions where selected if: (i) the minor allele frequency represented at least 5% of the total allele depth; (ii) had at least 100 reads of depth; and (iii) the minor allele nucleotides were supported by reads of both senses (at least 5% of depth should come from each read sense). In the running default, to detect iSNVs present at the minimum frequency of 5% an average depth of 2000 reads is required, for analysis with lower average depth the minimum depth threshold can be parsed.

The virus lineages signature is performed with Pangolin, the Pangolin and mutations constellation are updated at the moment of the Docker or Singularity images creation - to avoid outdated analysis - in interactive containers, the pangolin --update is strongly recommended. The consensus quality and set of mutations were evaluated with nextclade v.0.14.2. If the analyzed sample showed intrahost signals, the aforementioned analyses are performed for both consensus versions (with major and minor allele frequencies). On the other hand, the analysis will be performed only for the consensus genome with major iSNVs supported by nucleotides with major allele frequencies. In the last step, the assembly metrics such as depth and coverage were extracted with bamdst v.1.0.6 (https://github.com/shiquan/bamdst).

### Different environments, the same workflow

Considering the infrastructure and computational experience heterogeneity of different research groups working with SARS-CoV-2 genomic data we evaluated our workflow, and included documentation for two use cases (**Figure 1b**):

- Case I: Using a personal computer installing all dependencies; or using Docker or Singularity container services.
- Case II: Using a multi-user computational server, installing all dependencies; or using Docker or Singularity container services.

In this work, we simulate cases I and II. Case I was runned on a personal laptop with the following configurations: Ubuntu 20.04.2 LTS, 02 x RAM 4Gb DDR4 2133MHz and CPU Intel® Core™ i7-6500U 2.50GHz x 4, using 2 threads in the interactive Docker container. For Case II, we used a computational server with the following configurations: A node with 191Gb of RAM DRAM 2933MHz and two CPUs Intel(R) Xeon(R) Gold 5220R CPU @ 2.20GHz totalizing 96 threads, using 24 threads for the Singularity container runs. All command-line examples can be found in the documentation at the GitHub repository. Three independent runs were performed for each case.

### Data availability

The performance of the workflow was accessed using two datasets. The first one is a public dataset of 86 Brazilian SARS-CoV-2 Illumina paired-end libraries, generated by amplicon sequencing method using the Illumina COVIDSeq protocol, available under the EMBL-EBI study accession PRJEB47823, and the second one is an artificial dataset (**Supplementary file 1**) created with ART (Huang et al., 2012) of five paired-end libraries simulating a simultaneous infection (coinfection/codetection) of different SARS-CoV-2 lineages into a single sample (samples information on **Table S1**). The code and workflow documentation is available on https://github.com/dezordi/ViralFlow. And the supplementary data is available on [https://doi.org/10.6084/m9.figshare.16695262.v1].

## RESULTS AND DISCUSSION

Ninety-one genomic sequencing samples were assembled in both environments (see Methods, Case I and Case II). The total analysis running time - artificial samples are discarded of this estimation - in Case I was 9,38 hours (**Table 1**) with a mean time by sample equals to 6,7 minutes (stdev = 1,07, for details, see **Table S2**), while for Case II it took 3,06 hours, with a mean of 2,15 minutes (stdev = 0,17) for each sample. The use of RAM memory was 0,66 Gb and 0,73 Gb in Case I and Case II, respectively, this number was similar owing to the fact that all tools used in our workflow had their computational process based on CPU and not in RAM memory. A sequencing run with 96 samples (approximately 1.04Gb of throughput and 10,9 million reads) can be fully analyzed into 3,44 hours using a server with 24 threads or into 10,72 hours into a personal computer with 2 threads.

**Table 1.** Analysis time of a dataset with 91 samples with different computational structure.

| **Cases** | **RAM usage** | **Time per sample\*** | **Total time\*** |
| --- | --- | --- | --- |
| **Case I\*\*** | 0.66 | 2.15296 (0.17) | 183.60319 (0.022) |
| **Case II\*\*\*** | 0.73 | 6.734235 (1.07) | 563.98072 (7.11) |

\* Mean time and standard deviation in minutes; \*\* Personal computer; \*\*\* Computational server.

We detected a low number of iSNV, from 0 to 2, with a mean of 0 (stdev = 0.43, for details, see **Table S3**), for the eighty-six “non-artificial” samples supporting published estimates of low intrahost variant variability of SARS-CoV-2 (Naveca et al., 2021; Shen et al., 2020). For the five coinfection/codetection artificial samples, the workflow was able to consistently detect a large amount of well supported iSNVs (47 iSNVs per sample) (**Figure 2a, Table S4**). Those results show the capacity of the workflow to rapidly detect and generate a range of useful information that are important to generate new insights such as single consensus and coinfection of different SARS-CoV-2 lineages into a single sample. Moreover, the intrahost multi-allele frequencies can also be used to detect sample contamination, in a scenario where most samples show the same intrahost pattern, the more plausible explanation is sequence contamination rather than the origin of the same iSNVs in different samples.

**Figure 2.**
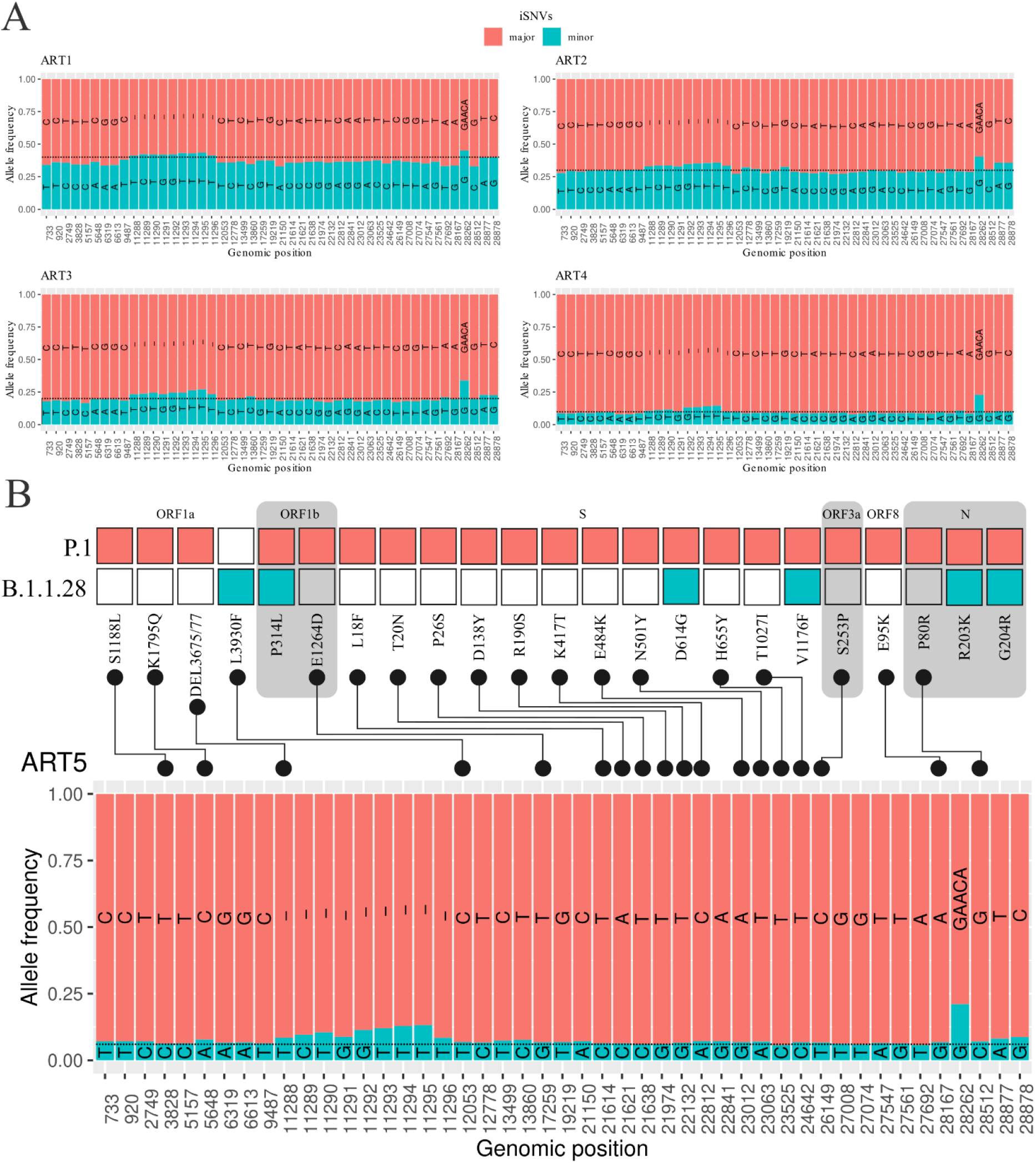
MAFs frequency sites of artificial datasets simulating co-infection events. The Black dashed line represents the expected minor iSNVs frequencies in each artificial dataset. **a**. The frequencies of 4 artificial datasets. **b**. The frequencies and associated mutations of minor and major consensus genomes.

Two key pieces of information are necessary for the deployment of SARS-CoV-2 outbreak control strategies: the virus lineage assignment and mutation characterization (Lauring & Hodcroft, 2021; van Oosterhout et al., 2021). The workflow generates two tabular files for each sample containing those key information: the ‘.pango.csv’ and the ‘.nextclade.csv’, which provide the virus lineage and the mutations found, respectively. The lineages identified in the 86 non-artificial samples (**Figure 3**) correspond to the set of mutations and to the expected lineages circulating at the collection date in each location (**Table S5**). For the 5 artificial samples, the virus lineages and set of mutations correspond to the allele frequencies present in iSNVs multi-allele frequencies (**Figure 2b, Table S5**). To show the precision of our workflow to detect the *indel* regions, 5 random non-artificial samples were assigned as P.1 with the deletion 11288-11297 into ORF1a and the insertion into the intergenic region at 28262 positions were manually curated with IGV (**Supplementary file 2**).

**Figure 3.**
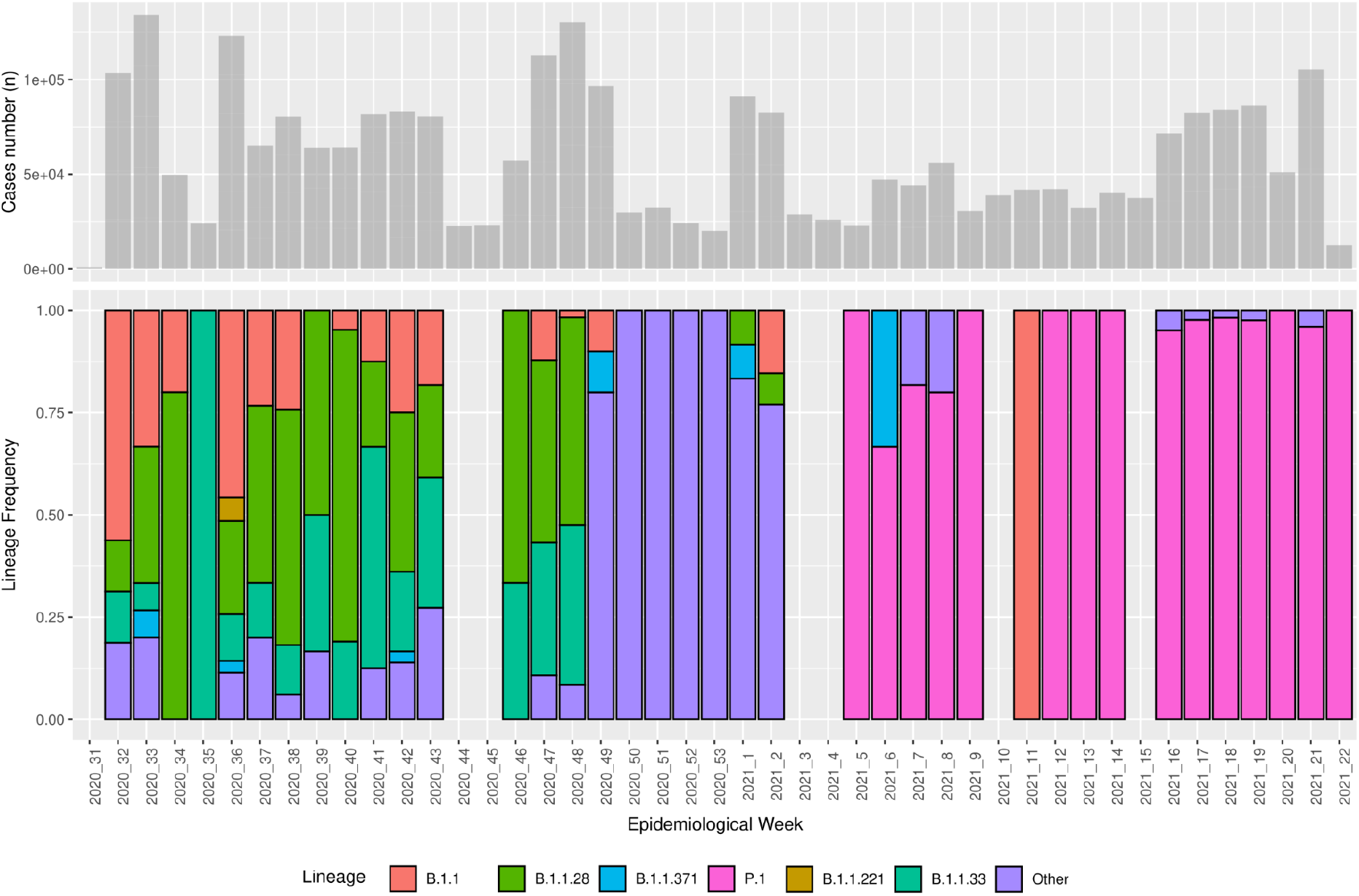
Lineages identified in consensus genomes with lineages found in the 86 samples dataset from Pernambuco state. Data recovered from GISAID at seplag.pe.gov.br at 30 Aug. 2021.

In addition to the intrahost, lineage, and mutations information generated by different tools present in our workflow, the ‘.quality.html’ generate by fastp and ‘coverage.report’ and ‘chromosomes.report’ files generated by bamdst that can be used for feedback to wet lab staff and to assess the quality of mapping and assembly steps. This information can also be crossed with the information of ‘qc.overallScore’ ‘qc.overallStatus’ columns present on ‘.nextclade.csv’ table.

Considering all the aforementioned results, the proposed workflow can be a good choice for groups that work with Illumina paired-end data and needs rapid information about both the quality of sequencing experiments, quality of consensus genome, and the lineage and mutation profiles that directly feed epidemiological reports.

## Data Availability

We used a public dataset of 86 Brazilian SARS-CoV-2 Illumina paired-end libraries, generated by amplicon sequencing method using the Illumina COVIDSeq protocol, available under the EMBL-EBI study accession PRJEB47823. The code and workflow documentation is available on https://github.com/dezordi/ViralFlow. And the supplementary data is available on [https://doi.org/10.6084/m9.figshare.16695262.v1].

## AUTHOR STATEMENTS

## Acknowledgments

We thank the LACEN-PE staff for providing the samples of SARS-CoV-2 and the Technological Platform Core for the support with their research facilities, the Fiocruz COVID-19 Genomic Surveillance Network for testing and source of information to compare results. We also thank all the researchers around the world that are working and generating data of SARS-CoV-2 in those difficult times.

## Author contributions

GLW and FZD conceived the study. GLW and TLC planned and supervised the work. FZD and GLW developed the workflow. FZD, PMCJ, CFA, SA tested and made improvements into workflow code. FZD carried out the tests, analysis and wrote the manuscript. TLC and FZD developed the containerization structures. All authors agreed with the final version of the manuscript.

## Ethical Aspects

This study was approved by the Aggeu Magalhaes Institute Ethical Committee—CAAE 32333120.4.0000.5190.

